# Near- and forecasting the SARS-COV-2 epidemic requires a global view and multiple methods

**DOI:** 10.1101/2020.04.09.20059402

**Authors:** Daren Austin

## Abstract

Conventional epidemiological models require estimates of important parameters including incubation time and case fatality rate that may be unavailable in the early stage of an epidemic. For the ongoing SARS-COV-2 epidemic, with no previous population exposure, alternative prediction methods less reliant on assumptions may prove more effective in the near-term. We present three methods used to provide early estimates of likely SARS-COV-2 epidemic progression. During the first stage of the epidemic, growth rate charts revealed the UK, Italy and Spain as outliers, with differentially increasing growth of deaths over cases. A novel data-driven time-series model was then used to near-cast 7-day future cases and deaths with much greater precision. Finally, an epidemio-statistical model was used to bridge from near-casting to forecasting the future course of the global epidemic. By applying *multiple* approaches to global SARS-COV-2 data, coupled with mixed-effects methods, countries further ahead in the epidemic provide valuable information for those behind. Using current daily global data, we note convergence in near-term predictions for Italy signifying an appropriate call on the future course of the global epidemic. For the UK and elsewhere, prediction of peak and eventual time to resolution is now possible.

## Introduction

Prediction of the ongoing global epidemic of SARS-COV-2 infections^1^ presents a number of challenges for epidemiology, notably the apparent susceptibility of the population at large, high transmissibility of the pathogen, lack of surveillance to estimate accurate incidence, prevalence and case fatality rate (CFR)^2,3,4,5^. Classic models of infectious disease transmission describe the processes using the law of mass action for contact transmission^6^. Refinements include mixing patterns, age stratification and spatial distribution. Whilst these models have proven extremely valuable for decision-making^7^, with validation using extended time-course datasets for multiple pathogens, they necessarily make strong parametric assumptions, from which prediction may be exponentially sensitive during the early growth phase of an epidemic. In this paper we present three complementary methods to inform on immediate, near-term (7-and 14-day) and further projection of the global SARS-COV-2 epidemic. Each method uses daily European Centre for Disease Prevention and Control (ECDC) numbers of new cases and deaths (ECDC)^8^, to inform at the individual country-level, the course of the ongoing SARS-COV-2 epidemic.

### Early projection using epidemic growth curves and doubling time ratios

During the very earliest stage of a new epidemic, where there is effectively unrestricted transmission into a new susceptible population, with little known about the underlying disease, an exponential model of the numbers of cases and deaths is appropriate. The number of cumulative cases, C(t) at time, t, is well-described using the simple exponential form, C(t) = C_0_ exp(r(t – t_0_)), with initial number of cases C_0_ at time t_0_, growth rate, r, incidence rC(t) new cases per day (also exponential) and doubling time log(2)/r days. The SARS-COV-2 infection has an incubation of approximately 5 days and a typical time from case presentation to death of approximately 14-21 days^4,5^. For any case fatality rate (CFR) and duration, T, between presenting as a case and eventual death, the number of deaths at a function of time is therefore D(t) = CFR C(t – T), which in the exponential phase is D(t) = CFR C_0_ exp(r(t – t_0_ – T)) = D_0_ exp(rt). Hence, we predict that for any (unknown) time delay and CFR, on a log-scale cases and deaths should initially have parallel slopes, with the distance between curves at any timepoint being defined as log(CFR). Deviations from parallel log-linearity (e.g., deaths out-pacing cases), may be informative as to when healthcare systems are beginning to fail. The reported CFR will vary from country to country, hence any global assessment must adjust for differences in surveillance practice.

The simplest prediction method is a direct comparison of any one country with the population of all other countries rebased to the same number of cases and deaths. Figure 1a shows this prediction for the United Kingdom from all data to 21/MAR/2020. Although prediction interval is wide, when every country’s cases and deaths were rebased to 64 and 4, respectively, early prediction of the UK epidemic was possible based on the same percentile growth charts familiar to any paediatrician^9^. The effects of global social distancing might be expected to move the trajectory of a country across percentiles, and this is suggested in more recent analyses (not shown).

**Figure 1.**
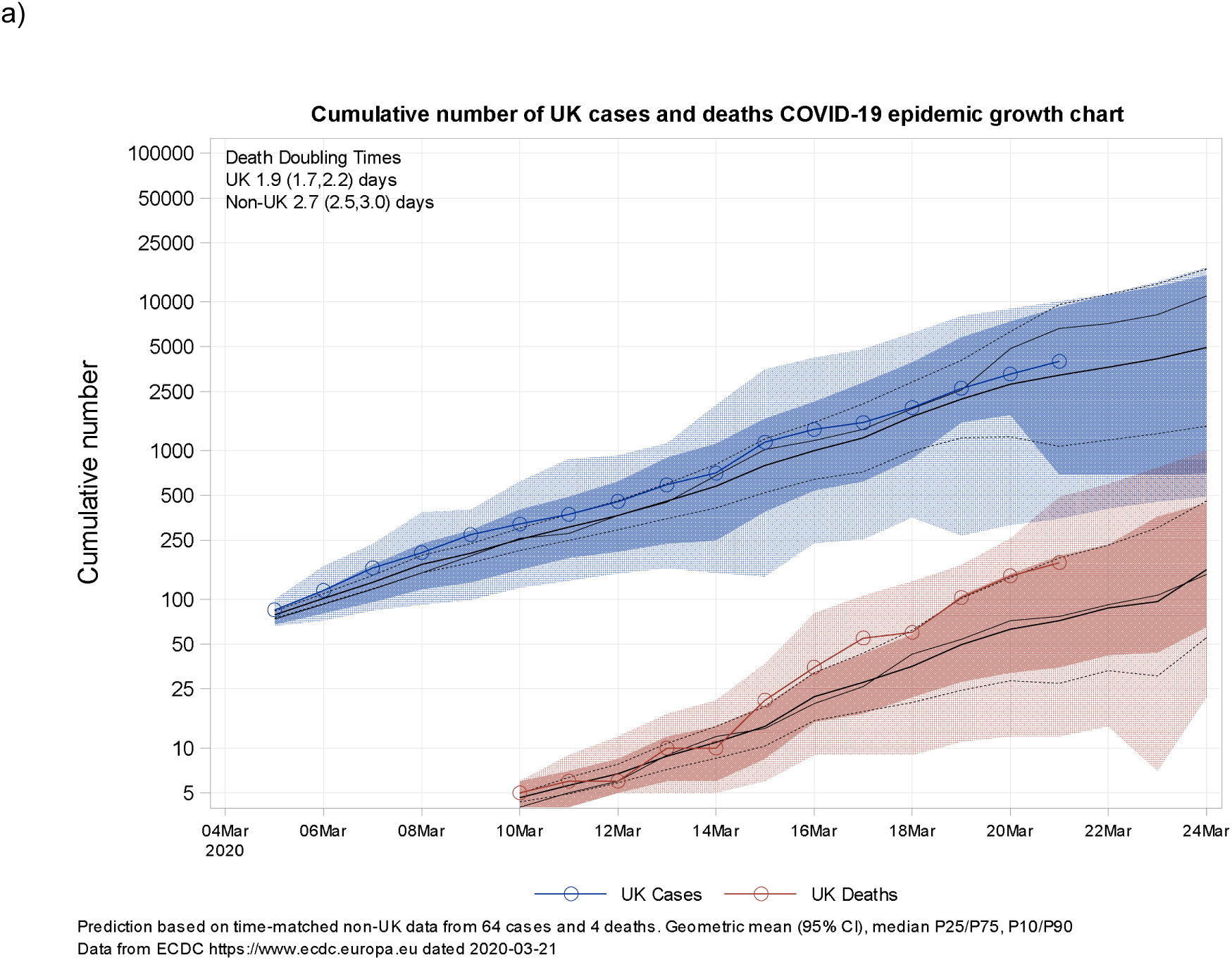
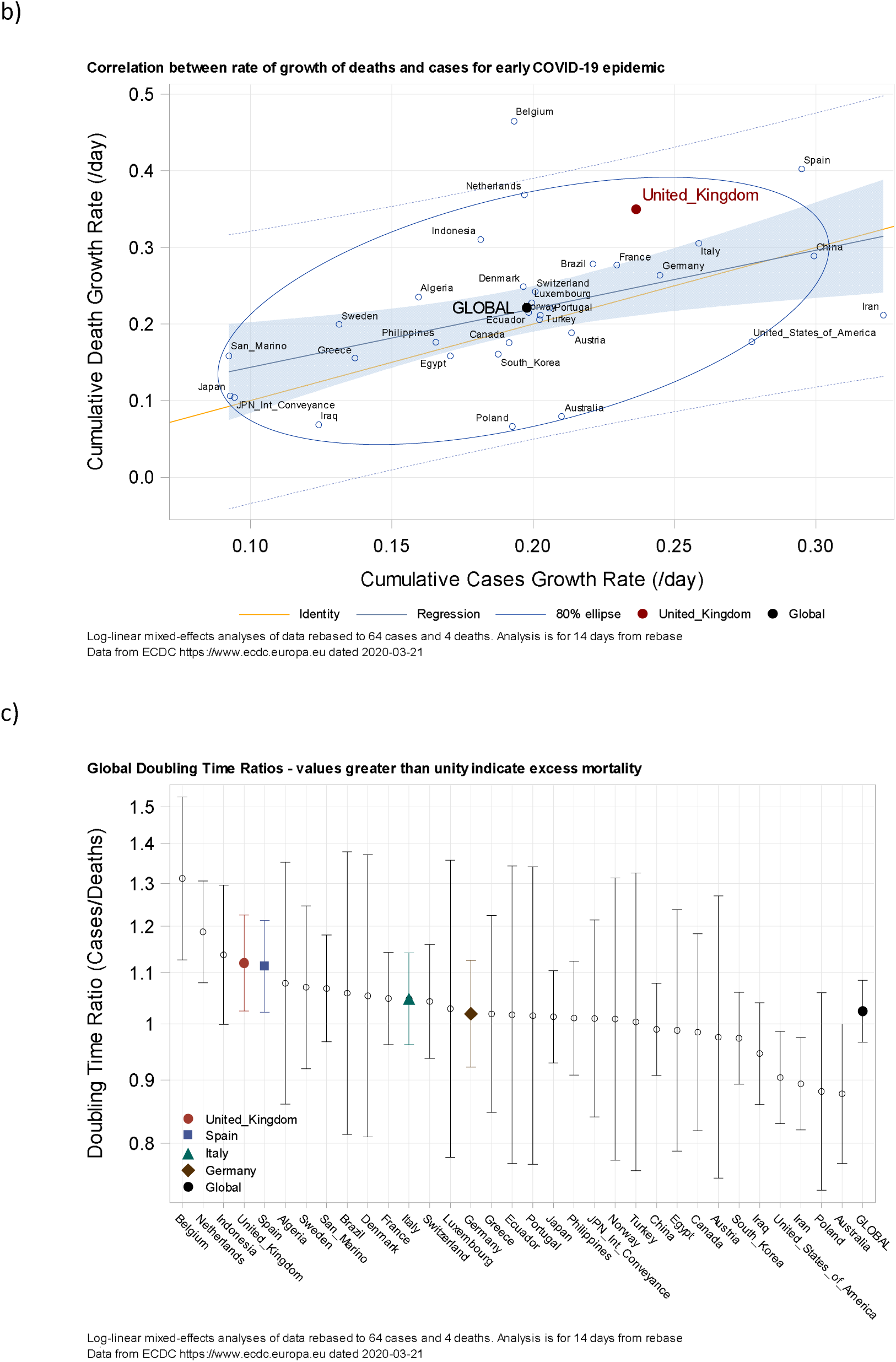
a) Growth curve for 2020 SARS-COV-2 global epidemic with UK data (not included in curve estimation) overlaid rebased to 64 cases and 4 deaths. Geometric mean and 95% CI, median, interquartile and interdecile ranges indicated. Data to 21/MAR/2020. b) Global exponential growth rates for cumulative cases and deaths with UK indicated (21/MAR/2020, data rebased to 64 cases and 4 deaths and fitted with a log-linear mixed effects model). c) Growth Rate Ratios for all countries, values greater than unity indicate that the rate of growth of deaths is greater than cases and may signify strains on healthcare (Data to 21/MAR/2020).

**Figure 2.**
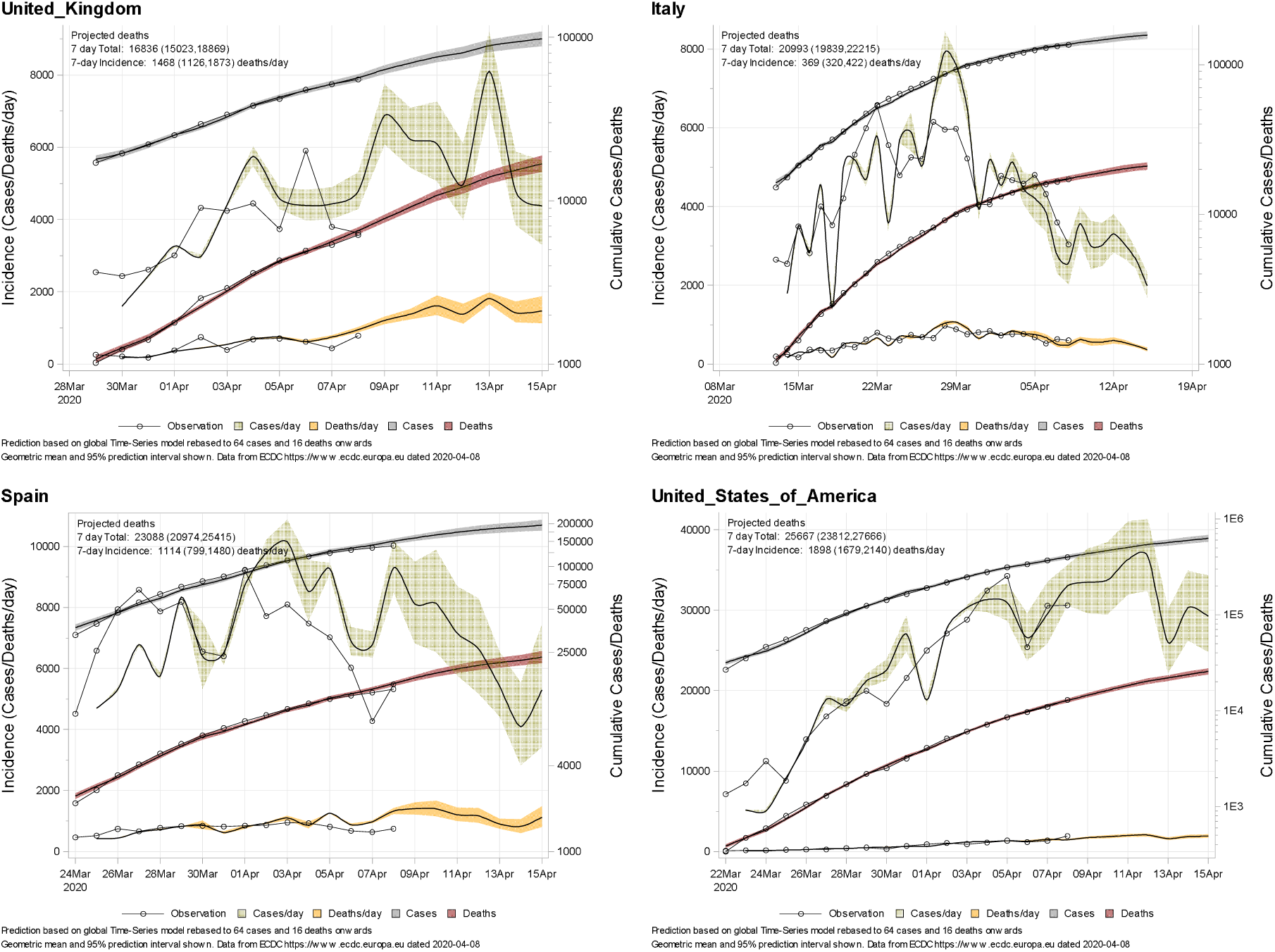
Near-term prediction of the ongoing UK SARS-COV-2 epidemic (mean and 95% prediction interval) for the dual-lagged linear mixed effects model. Model shows cumulative cases and deaths and daily case and mortality incidence. Near-casting (7-day) projection indicated. Data to 08/APR/2020.

A more sensitive analysis is provided using the Doubling Time Ratio (DTR), defined as the ratio of doubling times of cases compared to deaths. Figure 1b shows the correlation between exponential growth rates for cumulative cases and deaths estimated from log-linear mixed-effects models of global epidemic data (Date of analysis 21/MAR/2020), with their ratio shown (i.e., the DTR) plotted by descending DTR. The global mean exponential rate of growth of cases for the 33 countries and dependencies having reported 64 or more cases and 4 deaths, for the following 14-day period, was 0.20/day (95% CI 0.16, 0.23), doubling time 3.51 days (94% CI 3.01, 4.33), between-country variability 36%. The corresponding growth rate for deaths was 0.22/day (95% CI 0.17, 0.27), doubling time 3.15 days (94% CI 2.57, 4.08), between-country variability 52%. Increased variability in deaths reflects the early stage of the epidemic. Although the estimated global DTR was 1.02 (95% CI 0.97, 1.08), it is noteworthy that DTR for Italy and Spain, two EU countries particularly affected by COV-SARS-2, and the United Kingdom (DTR: 1.12, 95% CI 1.02, 1.23) were all significantly greater from unity. A DTR of greater than unity implies that deaths are outstripping cases (for any CFR), suggesting intervention measures are necessary to prevent further unrestricted differential exponential epidemic growth. An up-to-date analysis (Date 08/APR/2020) shows movement in UK DTR (1.07, 95% CI 1.04, 1.10) relative to the Global average (1.05, 95% CI 1.03, 1.06), suggestive of the effects of social distancing.

### Near-casting using time-series projection

Forward projection time-series methods have been used for many years in multiple disciplines^10^. Conventional time-series projections make assumptions regarding correlations in previous data to inform on future observations. Propagation of uncertainty, however, often limits precision (predictions are based on predictions not observations). The time-delay between cases and deaths inherent in the SARS-COV-2 epidemic lends itself to an alternative novel time-series methodology (Methods). Using past log-case data lagged to 7- and 14-days, and linear mixed-effects models we predict future cases based on past observations, and further predict deaths using the same model. All data is analysed in the log domain, with CFR described by a random effect to account for country-to-country variability in reporting practices. The use of dual lag times incorporates a (weekly) rate of change into the model. The minimum time delay of 7-days means that 7-day predictions are possible based on past *observations* (14-day predictions require one observation and one prediction, predictions beyond that use future predictions). This methodology opens a near-casting window on future events not available to conventional models, with the mixed-effects approach^11^ allowing prediction in countries that are still early in their epidemic.

Figure 2 shows sample predictions for selected countries (cut-off date 08/APR/2020, rebased to 64 cases and 16 deaths, 50 countries included in the analysis dataset) of cumulative cases, deaths and incidence. The model has a residual error of 11% (Extended Data) and is useful for near-casting of the SARS-COV-2 epidemic in all countries included, with sufficient precision and sensitivity (to 7-14-day time delays in actions) for helpful decision-making. The strength of this approach is the population-method used. Information from the global epidemic informs on individual country behaviour, and outlier behaviour can be identified early from individual country posterior model parameters (not shown).

### Long-term prediction using parametric epidemiological models

For long-term prediction, parametric epidemiological models will be necessary to control for exponential time-series prediction error. However, caution is advised on model selection and parameter estimation, most notably in the early epidemic phase. Conventional SEIR-type models assume a law of mass action to estimate case reproductive number, R_0_, which measures the transmissibility of the disease. Although intuitive, the notion of mass action, particularly in the presence of significant population-level social distancing (currently in place globally), may be limiting. Instead, we use the Gompertz model of epidemic growth, a limiting form of the logistic model, in which log-cumulative cases is described by the differential equation dlogC(t)/dt = r – slogC(t); r again being the unrestricted exponential growth rate, and r/s the eventual log-carrying capacity. The cumulative number of log-deaths is logD(t) = logCFR + logC(t – T), where the logCFR is a country-level case fatality rate, again modelled as a random effect to account for case reporting practices. The principal deviations of this model from conventional mass-action are the brake on epidemic growth (proportional to logC(t)) and log-carrying capacity. Transformation to the log-domain makes solution and model-fitting straightforward, controls for variability in highly non-linear processes and permits log-normally distributed random effects. The use of random effects describes country-to-country variability, particularly in surveillance and reporting methods, provided practices remain relatively consistent within a country during the epidemic.

Solution of the model for cumulative log(Cases) and log(Deaths) for country, i, with addition of 2018 population as a possible covariate in the model gives:

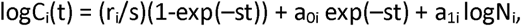

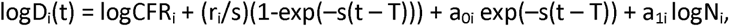

where (r_i_/s) is the log-final epidemic size (r_i_ being a random effect), a_0i_ the intercept (epidemic initiation, also a random effect), logN_i_ the reported 2018 World Bank population, and logCFR_i_ the individual country log-case fatality rate. The three random effects presume log-normal global distributions for epidemic initiation, eventual epidemic size and CFR.

Figure 3 shows results of the individual country predictions selected countries from analysis of global ECDC data (08/APR/2020 rebased to 128 cases and 32 deaths, 50 countries included in the analysis dataset). Overlaid is the time-series near-casting prediction for comparison and quality control. Where the epidemic shows significant curvature, estimation of peak incidence of death and overall epidemic size is possible. At the global level, population parameters are all well-estimated (Extended Data), without significant correlation for projection. The estimated global unrestricted epidemic growth rate is 0.69/day (95% CI 0.65, 0.73) – in a Gompertzian model, observed exponential growth is always less than r, between-country variability 73%, with CFR 6.3% (95% CI 4.9, 7.7), between-country variability 69%. Model residual for cases and deaths (modelled simultaneously with equal log-residual) is only 12%. Caution is necessary, however, regarding model interpretation – particularly eventual epidemic size, because model *selection* (as opposed to parameter estimation) may be invalid if unqualified by the near-casting model.

**Figure 3.**
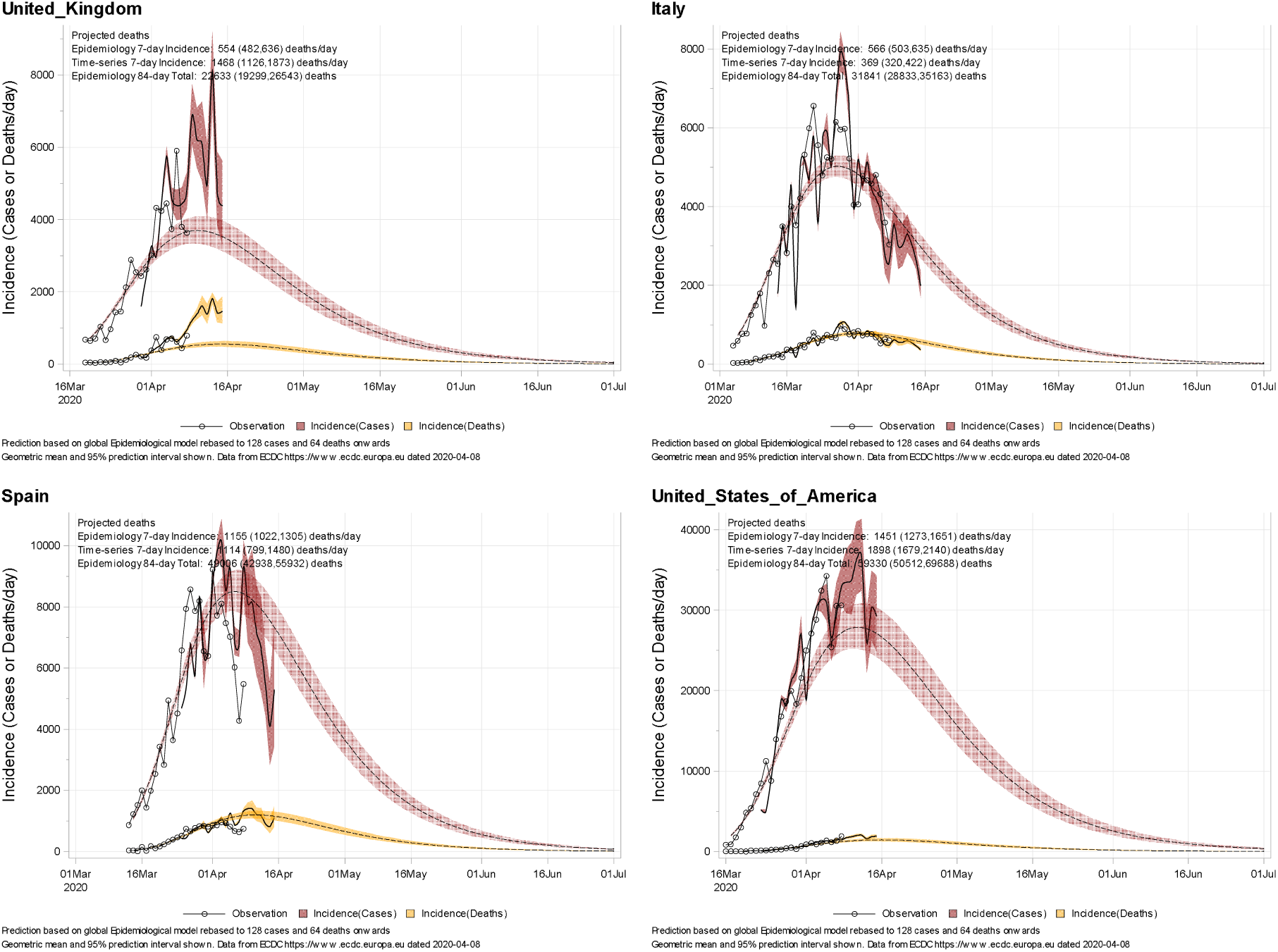
Posterior predicted course of the SARS-COV-2 epidemic for selected countries with near-term dual lagged model predictions overlaid. Concordance in short-term predictions improves confidence in the long-term projection Lack of concordance between prediction methods means that it is too early to call the final size and shape of the UK epidemic.

## Discussion

In the ongoing SARS-COV-2 epidemic, multiple methods are available for projection of numbers of cases and deaths. Without a means of projection, evaluation of the effectiveness (or otherwise) of intervention methods will prove challenging. Traditional epidemiology models based on contact and transmission patterns are important for planning intervention policies (e.g., mass vaccination and treatment), but they may struggle with calibration in a new setting when key parameters for their evaluation are poorly known. SARS-COV-2 presents such an example; an emergent global epidemic where previous immunity is absent. Robust intervention decisions will, however, be insensitive to parameter assumptions, and the predictions of Ferguson et al^7^, that there are no good outcomes without population intervention are therefore robust. In this analysis, we show that even in the absence of (key) epidemiological parameters (accurately known incubation time, underlying case fatality rate and transmissibility), by taking a global approach to the epidemic, it is possible to provide an early accurate assessment. We have shown that a simple “growth chart”, calculated from rebased case and death data from all countries other than the one of interest, was able to make an early prediction of the course of the UK SARS-COV-2 epidemic. This analysis, coupled with estimation of global log-linear growth rates for cases and deaths showed that the UK was likely to be an outlier, and that intervention was likely necessary. The Doubling Time Ratio for the UK showed that deaths were doubling 12% faster than cases, and faster than the global average, during the initial exponential phase (P<0.05 compared with a global estimate of equality), possibly implicating strain on healthcare services from an early stage of the epidemic. Similar findings were noted in both Italy and Spain.

Such a simple model is useful for immediate prediction but makes a strong assumption regarding future time course and is obviously unsuited to the first outbreak (in this case in Wuhan). It only assumes that progression (on a log scale) will follow other countries when rebased to the same point in the epidemic. Prediction intervals are necessarily very wide but still useful for the immediate (7-to 14-day time-frame) near-casting. Refinements such as scaling for population size (which was not available in the original ECDC dataset) may be unnecessary when an early epidemic is confined to the regional scale with population size of order 10^6^ (i.e., Wuhan Province, Northern Italy, London).

Once in the exponential growth phase, data-driven prediction is necessary. The characteristics of the epidemic (pseudo-exponential growth and an incubation from case presentation to eventual death) lend themselves to novel time-series analysis. We find that rebasing global data to a common number of cases, using 7- and 14-day lagged log-cumulative cases, and application of linear mixed-effects models (accounting for country-to-country variability), can be used to near-cast epidemic direction. The advantage over traditional time-series approaches is that the first 7-days of predictions are founded on past observations (not future predictions), thereby reducing (exponential) error. Case fatality rate differences were well-described at the country level by a log-linear random effect.

For eventual long-term forecasting, a parametric epidemiological model is necessary. Without such a model, a call on the location and size of the peak, and eventual size and duration of the epidemic, is not possible. These parametric models suffer from uncertainties and parameter correlations that make initial estimation and future very challenging from single country data. However, we find that using global, rather than individual country, data, a non-linear mixed-effects Gompertz epidemio-statistical model can describe the epidemic well with excellent parameter estimation and low residual. Providing a prediction of the time course for any single country from the model, however, requires some form of quality control. That quality control is provided by the near-casting model. Once near-term prediction intervals for the two modelling approaches coalesce, it is possible to move from near-casting to forecasting with confidence. Based on our most up-to-date analysis, this coalescence has happened in Italy, and it is now possible to provide more confident forward projections for that country and others.

The power of the epidemio-statistical global mixed-effects analysis is that, like the two previous approaches, countries further advanced in their epidemic inform on those following behind. We find concordance between the two approaches in Italy, with support for an eventual epidemic size of approximately 31.8k (95% CI 28.8k, 35.2k) deaths, with resolution by the beginning of July 2020. Using the model, we also find support for a peak in the UK epidemic somewhere from 7/APR/2020 to 21/APR/2020, approximately 22.6k (95% CI 12.3, 26.5k) deaths, and resolution also by July 2020. It is still too early to call the eventual global epidemic size, although values are predicted for every country in the analysis dataset. All predictions are founded on a continuation of the current global social distancing intervention program, which supports the use of a Gompertz epidemiological model. A relaxation of restrictions would release the brake on contacts, increase transmission, invalidate future model assumptions, and most-likely lead to an extension of the epidemic duration. Such predictions are out of scope for this analysis.

## Methods

Daily global data on new cases and deaths was downloaded from EDCD. Cumulative cases and deaths were log-transformed for analysis. Data was selected based on number of rebased cases, and time rebased from this point. Growth rate charts were calculated from geometric mean and percentiles assuming log-normal distribution, with doubling time estimated by log-linear regression. Doubling Time Ratio was estimated by separate log-linear random effects models, growth rates merged, and 95% confidence intervals calculated. For time-series projection, log-cumulative cases were lagged by 7- and 14-days prior to fitting a log-linear mixed-effects model with random intercept and slopes. Log-cumulative deaths was fitted to predicted number of cases, also with random intercept and slope. The epidemio-statistical Gompertz model was fitted to the same analysis dataset. Predictions (mean and 95% CI) were back-transformed, differenced (for incidence calculation) and overlaid with observation. Model goodness of fit was assessed visually and by Bayes Information Criteria. All analyses were conducted in SAS 9.4 proc mixed and proc nlmixed, automated by scripting and available from the author. At the time of publication, daily updated global projections are ongoing.

## Data Availability

All data and scripts are available from the author.

## Acknowledgements

The author would like to acknowledge the European Centre for Disease Prevention and Control, without who’s daily updated data, this work would not have been possible. This research is dedicated to all who have lost their lives to this dreadful disease.

## Extended data

### Time-Series model

Final parameters for linear mixed-effects Time-Series model of log(Cases); var1 = 7-day lagged log(Cases), var2 = 14-day lagged log(Cases). GeoID = Country ID.

| Solution for Fixed Effects |  |  |  |  |  |
| --- | --- | --- | --- | --- | --- |
| Effect | Estimate | Standard Error | DF | t Value | Pr > \|t\| |
| Intercept | 3.0981 | 0.4699 | 27 | 6.59 | <.0001 |
| var1 | 0.7920 | 0.08559 | 26 | 9.25 | <.0001 |
| var2 | -0.07047 | 0.04846 | 21 | -1.45 | 0.1607 |

| Covariance Parameter Estimates |  |  |
| --- | --- | --- |
| Cov Parm | Subject | Estimate |
| UN(1,1) | geold | 4.1598 |
| UN(2,1) | geold | -0.5955 |
| UN(2,2) | geold | 0.1002 |
| UN(3,1) | geold | 0.1834 |
| UN(3,2) | geold | -0.03779 |
| UN(3,3) | geold | 0.01943 |
| Residual |  | 0.002510 |

Final parameters for linear mixed-effects Time-Series model of log(Deaths); var1 = 7-day lagged log(Cases), var2 = 14-day lagged log(Cases).

| Covariance Parameter Estimates |  |  |
| --- | --- | --- |
| Cov Parm | Subject | Estimate |
| UN(1,1) | geold | 27.4397 |
| UN(2,1) | geold | -3.2088 |
| UN(2,2) | geold | 0.4123 |
| UN(3,1) | geold | 0.4031 |
| UN(3,2) | geold | -0.07932 |
| UN(3,3) | geold | 0.04143 |
| Residual |  | 0.002019 |

| Solution for Fixed Effects |  |  |  |  |  |  |  |  |
| --- | --- | --- | --- | --- | --- | --- | --- | --- |
| Effect | Estimate | Standard Error | DF | t Value | Pr > \|t\| | Alpha | Lower | Upper |
| Intercept | -1.5271 | 1.0530 | 27 | -1.45 | 0.1585 | 0.05 | -3.6876 | 0.6334 |
| var1 | 0.7129 | 0.1448 | 26 | 4.92 | <.0001 | 0.05 | 0.4152 | 1.0107 |
| var2 | 0.1953 | 0.06690 | 21 | 2.92 | 0.0082 | 0.05 | 0.05621 | 0.3345 |

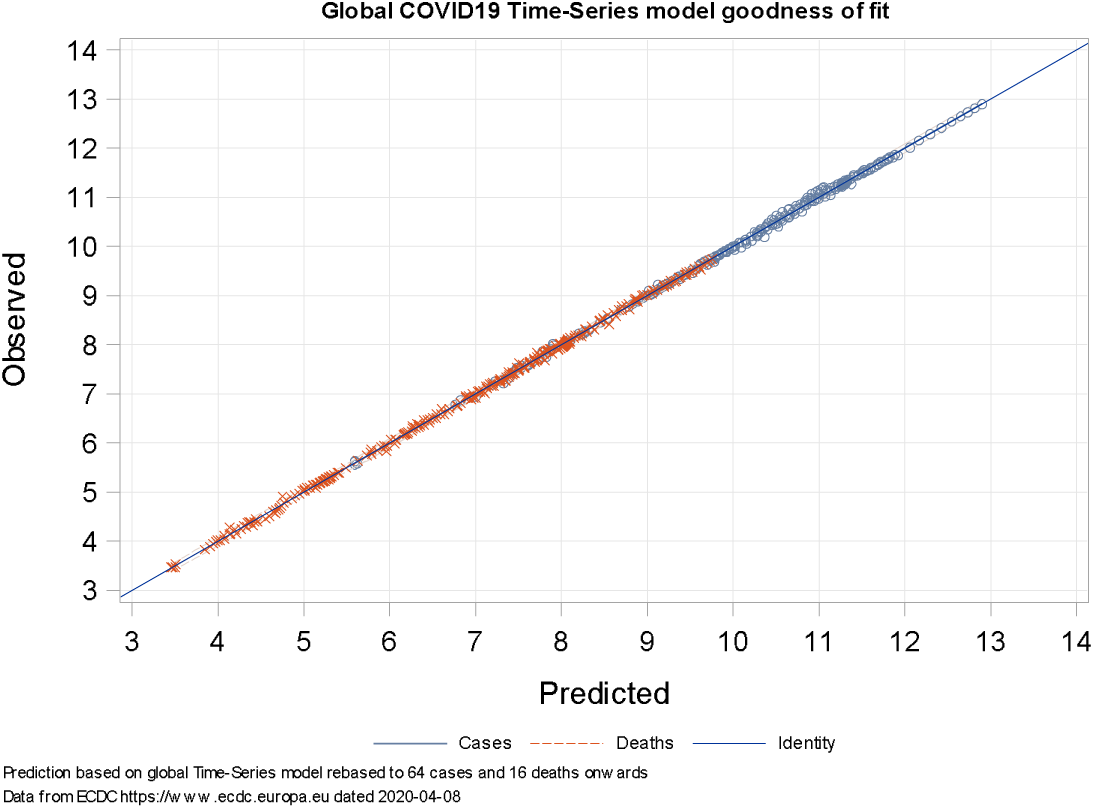

### Gompertzian epidemio-statistical model

Estimated model parameters and correlation matrix (population was excluded by BIC criteria)

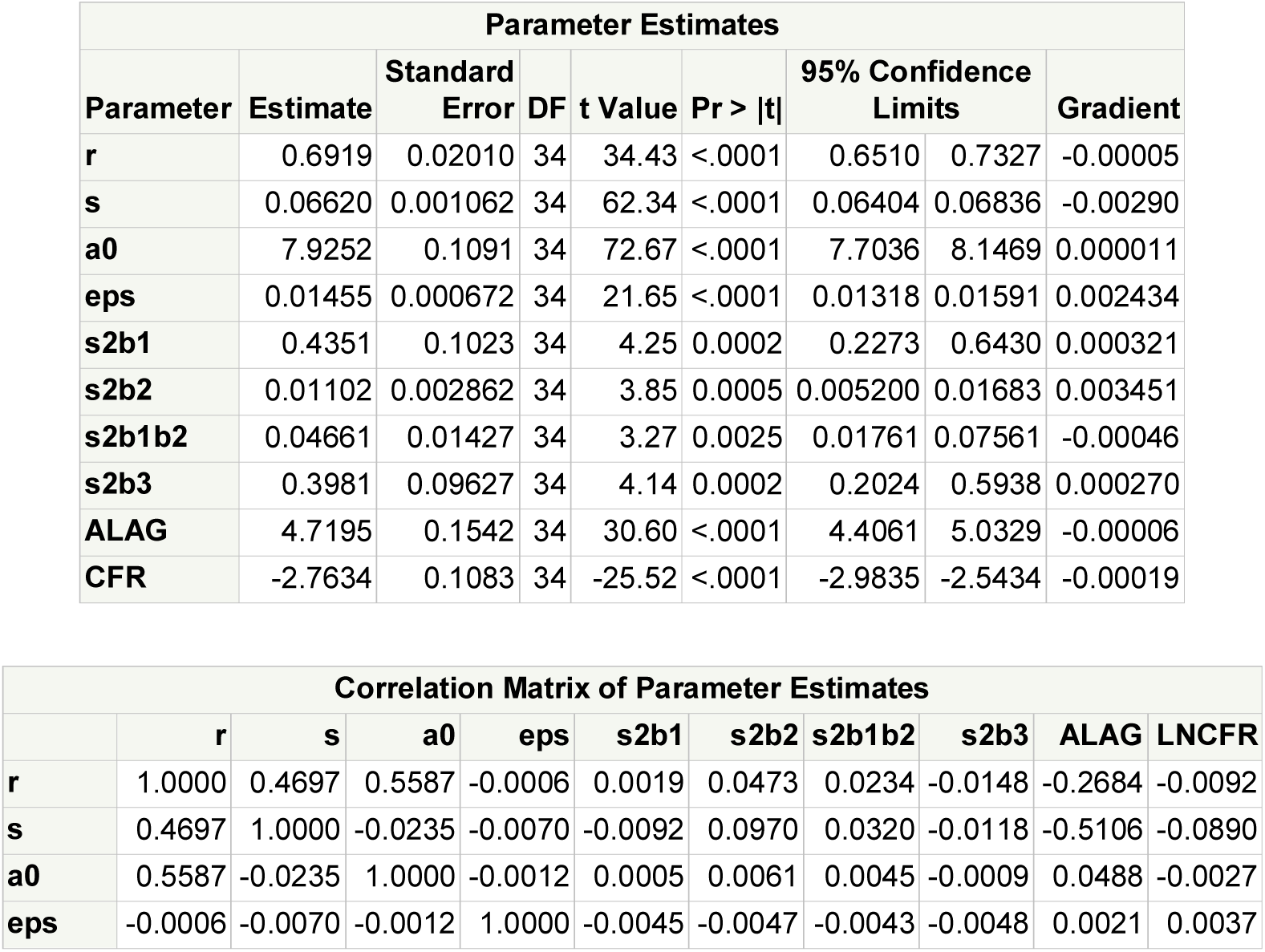
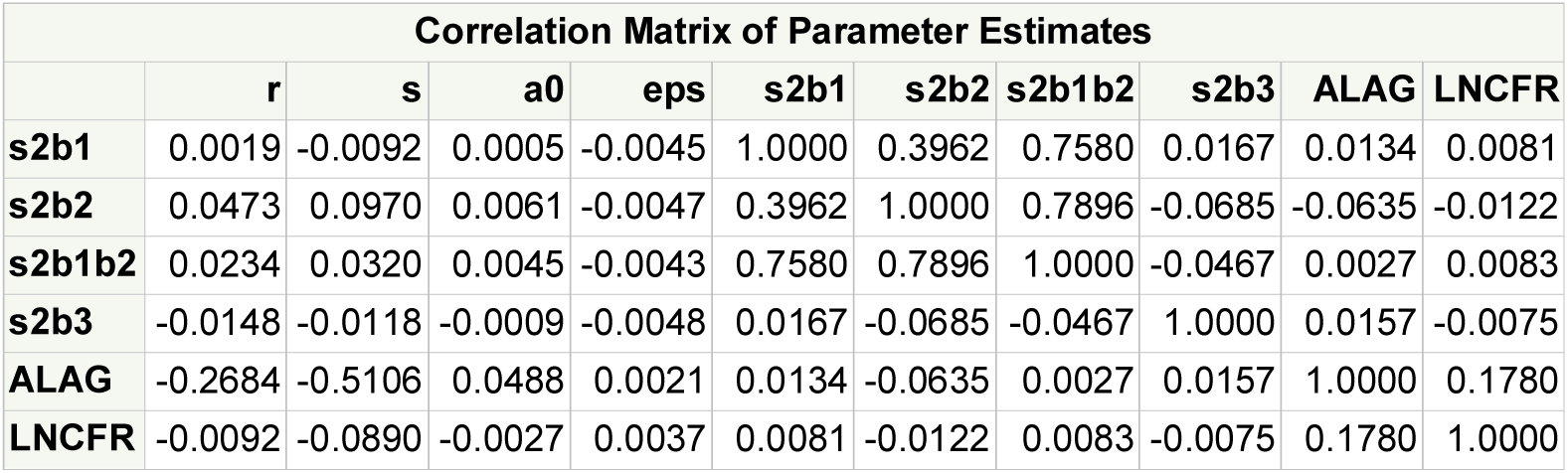

Parameters are: r= growth rate, r/s = log-carrying capacity, a0 = initial condition, eps = residual, s2b1 = variance(r), s2b2 = variance(a0), s2b3 = variance(logCFR), ALAG = lag time between cumulative cases and deaths, LNCFR = log(case fatality rate).

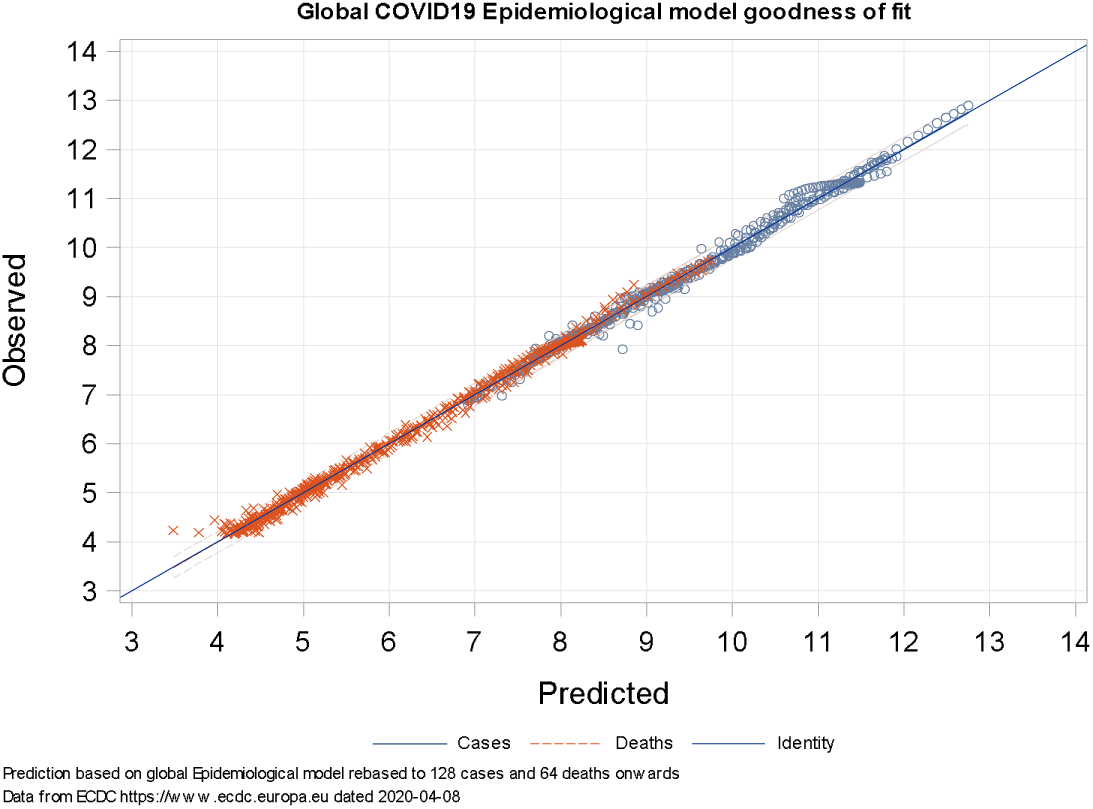

## References

1. World Health Organisation. (2019). Coronavirus disease (COVID-2019) situation reports. Retrieved from https://www.who.int/emergencies/diseases/novel-coronavirus-2019/situation-reports

2. Li, Q., & Wu, P. (2020). Early Transmission Dynamics in Wuhan, China, of Novel Coronavirus-Infected Pneumonia. N Engl J Med., 382(13), 382(13). doi:doi:10.1056/NEJMoa2001316

3. Li, R., Pei, S., Chen, B., Song, Y., Zhang, T., Yang, W., & Shaman, J. (2020). Substantial undocumented infection facilitates the rapid dissemination of novel coronavirus (SARS-CoV2). Science, eabb3221. doi:DOI: 10.1126/science.abb3221

4. Wang, D., Hu, B., & Hu, C. (2020). Clinical Characteristics of 138 Hospitalized Patients With 2019 Novel Coronavirus-Infected Pneumonia in Wuhan, China. JAMA, e201585. doi:doi:10.1001/jama.2020.1585

5. Zhou, F., Yu, T., & Du, R. (2020). Clinical course and risk factors for mortality of adult inpatients with COVID-19 in Wuhan, China: a retrospective cohort study. Lancet, 395(10229), 1054–1062. doi:doi:10.1016/S0140-6736(20)30566-3

6. Anderson, R. M., & May, R. M. (1992). Infectious Diseases of Humans: Dynamics and Control. Oxford: Oxford Science Publications.

7. Ferguson, N. M., Laydon, D., Nedjati-Gilani, G., & Imai, N. (2020). Impact of non-pharmaceutical interventions (NPIs) to reduce COVID19 mortality and healthcare demand. doi: DOI: https://doi.org/10.25561/77482

8. European Centre for Disease Prevention and Control. (2020). Publications and Data. Retrieved from https://www.ecdc.europa.eu/en/publications-data/download-todays-data-geographic-distribution-covid-19-cases-worldwide

9. Centers for Disease Prevention and Control. (2017). Clinical Growth Charts. Retrieved from https://www.cdc.gov/growthcharts/clinical_charts.htm

10. Montgomery, D. C., Jennings, C. L., & Kulahci, M. (2015). Introduction to Time Series Analysis and Forecasting, 2nd Edition. Hoboken: Wiley-Interscience.

11. Demidenko, E. (1987). Mixed Models: Theory and Applications with R, Second Edition. New York: Wiley Interscience. doi:10.1002/9781118651537

